# Study of Compliance of Law on Healthy and Balanced Diets by Schools in a District of New Delhi, India

**DOI:** 10.1101/2024.08.28.24312714

**Authors:** Raja Singh, Rima Dada, Arthur L Frank

## Abstract

There has been a steep increase in incidence of complex life style diseases. An unhealthy diet, alcohol consumption, tobacco consumption and high stress levels are major contributory factors. While action may be directed at dealing with all these points by various mechanisms, the issue of resolving unhealthy diets in schools, and the compliance of its related legal framework is discussed in this paper. In 2020, the Indian food regulatory organisation passed a regulation assuring school children are provided food that meets the standards of safety and a balanced diet. To this end, there has been consideration making sure foods with high trans/saturated fat and with added sodium and sugar do not form part of the diet of school children. All exposures to such foods, direct or indirect, have been prohibited and schools have been made accountable together with other local agencies. To understand compliance with this regulation, information from 49 public-run schools in a district of India’s capital New Delhi were collected and reported. The implementation and compliance status in these schools has been dismal with 24% being, for example, the maximum compliance for a single factor related to implementation of ‘Eat Right Campus’ campaign and similar or lower levels of compliance for other factors related to advertisement of unhealthy foods. The status of implementation and compliance of the food regulations for schools can inform policy and future actions required in this area. The creation of laws regarding regulation of schools must be followed by thorough implementation in order to deal with issues of rising global childhood obesity and other clinical manifestations (in the form of other lifestyle diseases) of unhealthy diets among children now and during adulthood.

## Introduction

In 2020, India notified the Food Safety and Standards (Safe food and balanced diets for children in school) Regulations, 2020^[1]^ under Section 92 of the Food Safety and Standards Act, 2006^[2]^. The original act in its intent states the act is ‘….to ensure availability of safe and wholesome food for human consumption.’ and Section 92 of the main Act deals with power of the Food and Standards Authority, also known as FSSAI to make regulations consistent with the act, especially related to ‘. (2) (e)notifying standards and guidelines in relation to articles of food meant for human consumption’ and for food safety and public health as in 92 (2) (g). These government regulations are intended to ‘promote the fundamental idea to make it clear what is healthy for children and what is not’ ^[3]^and it lays the responsibility in schools. This is also in line with the Indian jurisprudence which deals with a right to a healthy environment, like the need for good indoor air quality inside of schools ^[4]^.

The need for this regulation and other related issues was reported in a review study where 52 studies were analysed. The studies were from 16 of 23 Indian states and the study concluded that ‘overweight and obesity rates in children and adolescents’ were rising in the higher socio-economic group, in contrast to the double burden of having under weight children in lower income groups.^[5]^ Both obese and underweight children may show malnourishment. Similar results were also reported in a study from India’s capital, New Delhi, where 16,595 students were covered. It was reported that 12 percent of the students from Higher Income Group schools consumed fast food more than four times each week. The results from the lower income groups was 7.2% and from middle income group was 9.8%. The study reported overweight and obese students were present in the group aged 5-18 years^[6]^. The authors note that fast food is a highly inflammatory diet which is rich in trans fats/saturated fats, added sodium and sugars ^[7]^. The Lancet report states that unhealthy diet poses a greater risk to morbidity and mortality than does unsafe sex, alcohol, drugs and tobacco combined ^[8]^. This fast food diet not only leads to obesity and early onset of diabetes, hypertension amongst children, but it also adversely affects their mental health and well-being ^[9]^. This diet is also deficient in micronutrients and is associated with gut microbiome dysbiosis ^[10]^. A whole plant based diet, with seasonal fruits and vegetables not only nurtures the gut microbiome but also ensures its diversity, and is essential for mental health & well-being and optimal functioning of the immune system. Regular intake of fast food is associated with inflammation and switches on gene programs which are detrimental for health by impacting the epigenome. Another study from New Delhi, 1567 students of class 6th and 7th were selected from randomly selected private schools. 31.5% of these students were overweight.

The study recommended health-promoting interventions and healthy lifestyle information for the students. In this study, more than half of the students, irrespective of being underweight or overweight, mentioned that they eat junk food multiple times a week. The study also reported that 1/3rd of the participants had incorrect knowledge on basic nutrition and physical activity related behaviours ^[11]^.In another study from Delhi, where 1600 adolescents aged 10-19 years were analysed, ‘chi-square analysis revealed that consuming junk food and buying eatables from street shops was significantly associated to adolescents from age group 15-19 years’. This may point to the age where the exposure may be maximum. Further, faulty dietary habits like eating junk food and buying from street shops was associated with more participants being obese or overweight. ^[12]^Another study in New Delhi shows that the food and beverage options available in and around the participating educational institutes were high in one or more of fat, salt and sugar (HFSS). This is despite the institution guidelines that restrict the availability and accessibility of such HFSS foods. The vendors outside schools were present either during lunchtime or at the time of school closing, while around colleges the vendors were present throughout the day. The study also shows that HFSS advertisemenst were around private schools and colleges, but not around public schools. The study recommended stricter enforcement of the food regulations proximate to schools/colleges ^[13]^. It is often seen that there is availability of packaged foods like chips, biscuits etc, which have artificial flavours, colours and several additives and also products made from palm oil. A 2017 study from Kolkata, West Bengal, India showed that school canteens were advertising and supplying foods that were HFSS. This was in contrast with the message about healthy food that was given through instruction to the students. The study implied a need for healthy eating policies for schools. ^[14]^ This study was before the 2020 policy was made. There was one study from Bengaluru, Karnataka which specifically studied schools of urban slums. That study, in 2019, found that 27% of the 230 adolescents were overweight with 1.7% being obese. The link to junk food consumption and BMI variation was significant. ^[15]^ The regulation also pre-empts any evasive measures by schools which may outsource the responsibility to food contractors, but introducing the concept of FBO, or Food business operators, and making schools responsible for making sure these FBOs serve food that is ‘safe and balanced’ (Section 3[1])[1] There may be instances where the school would teach students to eat healthy diets, but on the other hand the school canteens would sell junk food. This is true not only in India, but has also been seen in the USA^[16]^

Despite the various studies that have been performed, no study has comprehensively looked at the full compliance of each and every legal requirement that the schools are required to be met. Most studies are based on an observational approach, where the actions taken by the school within may not have been systematically and comprehensively included. This is because the regulations for schools is not limited to mere prohibitory provisions of stopping the sale of foods high in saturated/trans fat or added sugar or sodium, but includes a comprehensive mechanism of positive inculcation of safe and balanced diets through various provisions which in summary include creation of health and wellness nodal teams, hiring of dieticians/nutritionists and among other methods creation of ‘Eat Right Campus’ through implementation of various provisions. Having such information may not only be required for understanding the status of legal compliance by schools, but is also useful for informing policy decisions with regards to food safety and the well being of school children. India, being a large consumer market with availability of consumer food products, may also need protection against childhood obesity which is a problem across the globe. ^[17]^ Parents should also be counselled to inculcate, as mentioned in the regulation, good healthy eating habits and ensure a balanced meal is given to their children. Children must also be discouraged to have junk food.

The detailed, point wise, list of compliances under the Food Safety and Standards (Safe food and balanced diets for children in schools) Regulations, 2020 are given in Table 1 below. These have been divided into basic category of compliance, namely, registration compliance, food selection compliance and advertisement and signages compliance. The advertisement is to include all form of surrogate advertisements that have been detailed in the regulations.

**Table 1:** Clause wise classification and description of the regulation with accountability details.

| S. No. | Compliance Category | Compliance Section | Compliance Details | Accountable agency | Enforcement to be done by: | Remarks |
| --- | --- | --- | --- | --- | --- | --- |
| 1 | Registration | 3 (1) | School authority selling or catering meals shall get registered | School Authority | School Authority |  |
| 2 | Registration | 3(2) | Food Business Operator selling or catering in school/having contract with school are to be registered | School Authority | Food Business Operators |  |
| 3 | Registration | 3(3) | All FBOs serving mid-day meal are to be registered | Department of School Education | Food Business Operators |  |
| 4 | FSSAI compliance | 3(4) | All FBOs shall ensure compliance to sanitary and hygienic practices valid for food manufacturers. | School Authority | FBOs | Sanitary and Hygienic practices as per Schedule 4, Food Safety Display Board and Food Safety Supervisors(s) as per Food |

|  |  |  |  |  |  |  |
| --- | --- | --- | --- | --- | --- | --- |
|  |  |  |  |  |  | Safety Standards(Licensing and Registration of Food Businesses) Regulations, 2011 |
| 5 | Food Selection/Compliance | 3(5) | Prohibition on sale of food products high in saturated fat/trans-fat or added sugar or sodium in school premises and campus | School Authority | School Authority |  |
| 6 | Advertisement (incl. Surrogate) & Signages | 3(6) | Boards containing warning 'Do not sell including free sale or market or advertise the food products high in saturated fat/trans-fat or added sodium or added sugar within school premises or campus' in English and one Indian language, to be displayed prominently at the entrance gate(s) of the school. | School Authority | School Authority |  |
| 7 | Advertisement (incl. Surrogate) & Signages | 3(7) | No advertisement (banner or wallpaper) of food which is high in saturated fat/trans-fat or added sugar or added sodium on school computers. | School authority | School authority |  |
| 8 | Food Selection/Compliance | 4(1) | Convert schools into 'Eat Right Campus' through comprehensive program promoting safe food and balanced diets with additional focus on local/seasonal food and no food waste. | Food Authority/State Food Authority | Schools |  |
| 9 | Food Selection/Compliance | 4(2) | Guidance from 'Dietary Guidelines for Indians-A manual' issued by National Institute of Nutrition and other expert institutions/guidelines to be followed for promotion of consumption of safe in balanced diet in school mess/canteen/kitchen. | School Authority | School Authority | Role of National Institute of Nutrition and expert agencies/institutions is highlighted. |
| 10 | Food | 4(3) | The prepared school | School | Food and | Role of the |
|  | Selection/Compliance |  | meals are selected based on the guidelines provided in the rules and as per food authority of the state and the commissioner of food safety of the state. | Authority | Beverage Operators (or contractors) | Commissioner of Food Safety and Food Authority for issuing guidelines. |
| 11 | Food Selection/Compliance | 4(4) | Nutritionists, dieticians, nutrition associations or parental help may be sought for drafting the menu for the children at regular intervals. | School Authority | School Authority | The importance of taking parents into the loop is highlighted. |
| 12 | Food Selection/Compliance | 4(5) | The creches/day cares for infants and children upto 2 years old are also expected to serve safe and balanced diets. | State Food Authority | Creches/ Day cares for infants and children upto two years | The use of word 'expected' makes it non mandatory and discretionary. |
| 13 | Advertisement (incl. Surrogate) & Signages | 5(1) | Prohibition of advertisement/selling/market/offer for free or sale/permit sale of food products high in saturated fat/trans-fat or added sugar or sodium in school campus or to school children in an area within fifty metres from the school gate in any direction |  |  | Responsibility not explicitly defined |
| 14 | Advertisement (incl. Surrogate) & Signages | 5(2) | a, During marketing of foods, no incentive shall be given for foods which is high in saturated fat/trans-fat or added sugar or sodium.<br>b, no sponsorship shall be taken for food high in saturated fat/trans-fat or added sugar or sodium | School Authority | Food and Beverage operators | Premiums and incentives include toys, trading cards, apparel, club memberships, contests, reduced-price specials, or coupons with foods, meals.<br>Sponsorship includes of sporting, school, and other events for children. |
| 15 | Advertisement | 5(3) | No food with | School | Food and |  |
|  | (incl. Surrogate) & Signages |  | saturated/trans fat or added sugar or sodium shall be marketed, sold or given away in form of advertisement, or educational incentives, free samples, or fundraising activities. | Authority | Beverages operator |  |
| 16 | Food Selection/Compliance | 6(1) | A 'Health and Wellness Ambassador' and a 'Health and Wellness Team' is to be appointed which shall regularly perform inspections in schools. This shall also monitor, as nodal persons, availability of safe, balanced and hygienic food. | School Authority | Health and Wellness Ambassador and Health and Wellness Team |  |
| 17 | Food Selection/Compliance | 6(2) | Regular surveillance and periodic inspections of food business operators shall be performed. | State Food Authority | Food and Beverage Operators | Compliance of Act, rules and regulations is to be done. |
| 18 | Food Selection/Compliance | 6(3) | The compliance related to promotion and sale as mention in Section 5 shall also be done by public authorities like the municipal authorities/local bodies/panchayats as notified | Municipal Corporations, local bodies, panchayats |  |  |
| 19 | Food Selection/Compliance | 6(4) | Action when violation of rules: The state food authority can take up the matter against the school to the department of school education and affiliation bodies. | State food authority | School authorities | Department of School Education/ Affiliation bodies like Central Board of Secondary education are to be involved. |
| 20 | Food Selection/Compliance | 6(5) and 6(6) | The State Legal Advisory Committee shall create a subcommittee to monitor implementations of these regulations. |  | Sub Committee shall consist of Department of School education, public health professionals in filed of food and nutrition. | This is under the Sub regulation 2.1.15 of the Food Safety and Standards (Licensing and Registration of Food businesses) Regulations, 2011. To meet twice an year. |
| 22 | Food Selection/Compliance | 6(7) | The indicative list of local food groups and preparation shall be made by the sub committee as per the broad guidelines. |  |  |  |
| 23 | Food Selection/Compliance | Schedule | General guidelines and selection of food to be given, |  |  |  |

## Aims and Objectives

There is need for a study which comprehensively covers the compliance of the food regulations. In order to cover this gap for a study inclusive of positive efforts by schools and a comprehensive analysis of compliance by schools, the current study was undertaken. The aim of the study, as stated above, is to check whether government schools in a district of New Delhi comply with the provisions of the Food Safety and Standards (Safe food and balanced diets for children in schools) Regulations, 2020 with respect to the major points for which the school may be directly responsible.

## Materials and Methods

In order to perform the study of the compliance to the comprehensive provision of the food regulation for school, the following steps were taken:

1. The need for the study has been established through a brief literature review.
2. The aims and objectives have been stated.
3. A comprehensive list of information sought from schools, where schools are responsible for compliance related to the balanced and safe diet regulation is made.
4. The requests for information under the Right to Information Act 2005 are sent to schools and the responses from schools are collected.
5. The information provided by schools is analysed and reported.

The full detail of information collected from schools is listed in table 2 below.

**Table 2:** The Information that was sought from the schools to check for compliance of the regulation.

| S. No. | Information Sought | Remarks |
| --- | --- | --- |
| 1 | Information related to the contract with any food business operator selling or catering food in the school campus, in case the school is not into the sale |  |

|  |  |  |
| --- | --- | --- |
|  | of such food. |  |
| 2 | Number of instances recorded where the school has prohibited the sale of food products high in saturated fat or trans-fat or added sugar or sodium in school premises or campus as per Section 3(5) of the Food Safety and Standards (Safe food and balanced diets for children in schools) Regulations, 2020 | The compliance lies with the school to implement the regulations. |
| 3 | Record of the board as per Section 3 (6) of the Food Safety and Standards (Safe food and balanced diets for children in schools) Regulations, 2020, which has been placed in the school along with location and text of the board. | The board is to be placed at the entrance of the school. |
| 4 | Details of any advertisement containing foods high in saturated fat or trans-fat or added sugar or added sodium. |  |
| 5 | Details of steps taken by the school to implement 'Eat Right Campus' as per Section 4(1) of the Food Safety and Standards (Safe food and balanced sites for children in schools) Regulations, 2020 | This is a positive compliance where the school becomes the spreader of information about health diet. |
| 6 | Details of the menu that is served in the school canteen, school mess or any other place with any other name where food is sold or served to the students. |  |
| 7 | Details of any nutritionists/dieticians hired by schools as per Section 4(4) of the Food Safety and Standards (Safe food and balanced diets for children in schools) Regulations, 2020 |  |
| 8 | Details of the Health and Wellness Ambassador and the Wellness Team as per Section 6 (1) of the Food Safety and Standards (Safe food and balanced diets for children in schools) Regulations, 2020. | The team and the ambassador are responsible for regular inspections. |
| 9 | Details by name and designation, with contact details of the School Authority as per Section 2(1) (i) of the Food Safety and Standards (Safe food and balanced diets fo children in schools ) Regulations, 2020 |  |
| 10 | Details of how school identifies itself as per Section 2 (1) (h) of the Food Safety and Standards (Safe food and balanced diets for children in schools) Regulations, 2020 |  |

The period of collection of information for this study is the later part of 2023. The limitations in this study are namely, the non inclusion of private schools. This is because the study has utilised the Right to Information Act 2005, which is empowered only to deal with public authorities which includes only government schools. Secondly, the study takes the information provided by schools directly and this has not been further validated. This is because the schools have provided the information under their seal as part of a legal requirement and are less likely to provide incorrect information as the accountability lies with the school for the information provided. Thirdly, the study has been performed only in one district of New Delhi. Though 49 schools have been covered, it may not tell the real picture for the complete city, state of the country. Despite the above limitations reported, the study provides insight into 49 government schools in the North District of Delhi. This may be used for informing policy in the area of food safety and in steps dealing with childhood nutrition and obesity in the country.

The study does not involve any human participants, drugs, animals or human tissue. Further, the study uses information that has been released as information in open public domain by the schools. The study does not collect any personal information from the school about any children or any human participant. By virtue of the data released under the Right to Information Act 2005, no personal data can be released by any public authority ^[18]^. The names of the schools have also been anonymised. The study deals with compliance of law related to food. Due to the above, the study does not fall under the purview of ethics review and the authors declare the same.

## Results and Analysis

On the first issue of a contract with food and beverages operators with respect to selling or catering food in the school campus, out of 49 schools, only two stated that they followed the guidelines from directorate of education as far as mid day meals are concerned, while one stated that there was no canteen in the school. The rest either stated that it was not applicable, or did not pertain to the school or was the responsibility of the directorate of education.

The second was the issue of the number of instances recorded where the school has prohibited the sale of food products high in saturated fat or trans-fat or added sugar or sodium in school premises or campus as per Section 3(5) of the Food Safety and Standards (Safe food and balanced diets for children in schools) Regulations, 2020. Out of the 49 schools, only two stated that there was no such case in the school, while one stated that the school only provided mid day meal and hence was not selling or providing any other food product. The 46 other schools either stated that this either did not pertain to them or was not applicable to the schools.

The third was the issue of the record of the board as per Section 3 (6) of the Food Safety and Standards (Safe food and balanced diets for children in schools) Regulations, 2020, which has been placed in the school along with location and text of the board. Out of 49 schools, only six stated having the boards installed at the entrance as per requirements. Two stated having charts within the school in classes or on the grounds. One did not provide information about the boards but stated that students are guided verbally during break times. The remaining 40 schools denied information by stating that either it was not applicable to them or not in their purview.

The fourth was the issue of details of any advertisement containing foods high in saturated fat or trans-fat or added sugar or added sodium. Only one school mentioned the foods for which advertisement was present and action may be taken. One other school mentioned that students were briefed in the assembly. The remaining 47 schools did not provided information and stated that this was either not applicable or was under the preview of the department of education.

The fifth was the details of steps taken by the school to implement ‘Eat Right Campus’ as per Section 4(1) of the Food Safety and Standards (Safe food and balanced sites for children in schools) Regulations, 2020. Out of the 49, only 12 schools stated following some measures related to the ‘Eat Right Campus’. Out of these 12, two provided full detailed steps, with the rest 10 stating common measures like sensitisation in morning assembly.

The sixth was the details of the menu that is served in the school canteen, school mess or any other place with any other name where food is sold or served to the students. 5 schools provided the details of the menu of the schools. There were others which stated that there was no mess or canteen in the school, but the issue of service of mid day meal was not addressed by those.

The seventh was the details of any nutritionists/dieticians hired by schools as per Section 4(4) of the Food Safety and Standards (Safe food and balanced diets for children in schools) Regulations, 2020. On this five schools stated that no nutritionist/dieticians were appointed. Four schools out of 49 stated that teachers (of home science subject) in particular provided counselling.

The eighth was the details of the Health and Wellness Ambassador and the Wellness Team as per Section 6 (1) of the Food Safety and Standards (Safe food and balanced diets for children in schools) Regulations, 2020. Nine schools had a person responsible with three schools specifically providing details of health and wellness ambassador.

The ninth and tenth issues were regarding details of school and nomenclature of the school and was not particularly important for this research but was for record keeping.

It is clear from the above information that the implementation and compliance of the food regulation inside and around the schools is far from universal and not fully complied with. It must be stated here that one school in Model Town, Delhi (name anonymised) was the most responsive and aware about the provisions with compliance to many points of the regulation.

## Discussion

This paper notes the compliance status of regulations made to provide steps in making sure schools provide safe and balanced diets to the students. In the case of the one district of New Delhi, the implementation and compliance is dismal. The regulation provides a very comprehensive coverage of all points inside and outside of the schools that may be required for the Indian scenario. Clearly, there has been little oversight of the implementation required by the regulations.

India is in a vulnerable position as there is a double burden from infectious diseases as well as from non-communicable disease. Instead of looking at these as binary, resource allocation should be there for communicable as well as non-communicable diseases. These non communicable diseases included diabetes, ischemic heart diseases, chronic obstructive pulmonary diseases and cerebrovascular diseases. ^[19]^ There has also been an increase in the cases of Polycystic Ovarian Syndrome, which has increased in both urban and rural setups, due to junk food, sedentary lifestyle, binge-eating and easy availability of nutritionally deplete carbonated drinks ^[20]^. The role of optimising nutrition for better physical and mental health cannot be ignored ^[9]^. One of the major factors for a steep increase in mental health disorders like Attention Deficit Hyperactivity Syndrome, autism, bipolar disorder and disability due to mental health issues ^[21]^is due to poor nutrition and over consumption of highly processed food, a diet rich in trans fats, salt, sugar and refined grains and artificial sweeteners. This diet is highly inflammatory and lacks vital nutrients and micronutrients. A healthy, nutritious, whole plant-based diet nurtures the gut microbiome and ensures its diversity ^[22]^. A healthy gut microbiome is necessary as the bacteria secrete various factors important for mental health and also one important for our immune system ^[23]^. Gut microbiome dysbiosis is one of the factors leading to lifestyle diseases ^[24]^. Our lifestyle, thoughts, eating habits also influence our epigenome, and this may have an multi generational effect among populations ^[25]^.

Early onset diabetes is increasing in India^[26]^. There is also early onset diabetes among youth with it being called an ‘awakening epidemic’ ^[27]^ One way to deal with this is to regulate the food choices for populations, especially early in life. The policy to regulate this^[28]^ is important especially due to a rise in the autonomy that adolescents have in food choices over time. ^[29]^ Schools play a key role in these food choices as children spend substantial time away from direct parent supervision and are influenced by multiple factors such as commercial sale of food within and outside of school. ^[30]^ There are also advertisement factors that influence buying decisions of school children outside and within the school. ^[31]^ There may be advertisers that sponsor activities in school and these are sellers of food with high trans/saturated fat and added sugar or sodium that may be subconsciously promoted through advertisement in these school. The regulation checks most of the factors and is comprehensive in nature, but appears to be poorly enforced.

With respect to early onset childhood obesity in India apart from interventions like increased physical activity, reducing screen time, the role of schools in providing awareness and education is important, including regarding diet. ^[32]^ There is a need to make parents aware as they have the capability of providing nutritional food in tiffin and discuss the issue of compliance of food regulation with the school authorities.

Further, the link to obesity may depend on many issues like public vs private schools, rural vs urban area and the socio economic situation of the area where the schools are located. But in any case, there is a need for better oversight regarding the regulation of foods that are served inside and outside of the schools. ^[33]^ and the ‘Eat Right Campus’, campaign is an important step in the right direction. But as shown in this study, the implementation needs more vigour. It has also been reported that this campaign initiated by the FSSAI, or the Food Safety and Standards Authority of Indian has not received a very positive response and more institutions need to become certified. ^[34]^

Regarding the choice of menu, there has been a very detailed classification of the suggested menu in the regulations which prescribes a general guideline for selection of foods. This includes division of foods into three categories as follows:

1. Always a part of the menu-75-80% of the foods should come from this category. This includes cereals (millets and pulses included), milk products, fruits/vegetables and healthy nuts and cooked foods which are cooked in a healthy way.
2. Those be eaten occasionally in small portion size and reduced frequency (eg once per week). This includes deserts, packed foods, bakery products and some beverages.
3. Not to be made available in the school, hostel, etc. This includes food products high in saturated fat/trans fat or food that may have added sugar or sodium. ^[1]^

For the above to be implemented, high levels of awareness and oversight will be needed by school authorities, parents and children so that they can also themselves make informed choices. The step to designate health and wellness ambassadors and teams is in the right direction, but needs thorough implementation, which this study reported as not universal. There is also a lack of clarity in the law about accountability. Even though, in Delhi, the Directorate of Education has issued mandatory directions for all schools to follow, there seems to be an absence of regular follow up, as the results of this study shows. ^[35]^

## Recommendations

Firstly, parents and children need to be educated about role of diet in health, the need to have home cooked balanced meal as outside food may compromise quality for taste. Thus, only healthy food should be made available with an emphasis on seasonal fruits and vegetables and whole grains. We also emphasise that a 30 min to 1 hour sessions per month should be dedicated to educating children on importance of a healthy balanced diet and during PTM parents also need to counselled and educated.

Secondly, changes in the lifestyle can also include Yoga, which is easy to perform, requires no equipment and is free from the compulsions of outdoor or indoor preferences. Yoga promotes health, prevents onset of diseases and can be used as adjunct management of diseases. Yoga comprises mostly *asana, pranayaam* and *dhyana,* but also advocates a whole plant-based diet with intake of fruits and vegetables. This practice can improve both the physical and mental health of children. Yoga is an ‘art of living’ and this with a whole plant-based diet is an integral part of the lifestyle. This can enhance children’s concentration, elevate memory and emotional resilience, and also regulate their sleep-wake cycle. A fast food diet also influences our germ cells and thus it can determine trans-genetic effects. Studies from the lab of the second author has documented that Yoga truly moderates the epigenomes and switches on the ‘internal pharmacy’ by switching on gene programs beneficial for health (the antioxidants, anti-inflammatory genes maintaining gut integrity, increased DNA repair genes, genes maintaining nemoplasticity^[36][37][38][39]^.

Thirdly, to make sure the well thought out regulation does not remain un-implemented, the accountability of the schools and the other agencies involved must be brought under a tighter control. Incentives and punishments for compliance or lack of it, respectively, should be pursued on an urgent basis.

Fourthly, like in the case of pulse polis programs, AIDS control program and many such national public programs, the use of IEC (Information, Education and Communication) techniques like movies, advertisements and posters should be developed. These must include celebrities and be delivered with high quality ^[40]^. Corporates should be encouraged to provide IEC materials, as most schools lacked a basic board for stopping of sale of HFSS food.

Fifthly, it was noticed that public sector schools did not have access to nutritionists and teachers had taken over the work of awareness training. In order to truly implement the food regulations in schools, a cadre of nutritionists and dieticians must be employed in public schools in order to have a professional approach for implementation of the food regulations in schools.

Sixth, for the implementation of the stoppage of sale of HFSS food around the schools, the local police should assist the school authorities as the school authorities may be subjected to intimidation by a nexus of food sellers. To assist the police, civic volunteers like Delhi Home Guards or other such human resources may be utilised.

Further, the issue of health of school children, apart from healthy diet and its regulation around the school, also needs the strict implementation of the tobacco control law. It has been found in previous studies that many schools in New Delhi do not fully comply with the tobacco control regulations ^[41]^. There are also concerns regarding good indoor quality^[42]^ in buildings like school buildings and issues like the use of asbestos^[43]^. While indoor air quality and use of asbestos needs a top-down approach of policy intervention followed by executive will, putting signboards, which is an easy method of compliance but absent in many schools, needs decentralised effort by school authorities.

## Conclusion

The aims of the study was to highlight the need of a good/balanced and nutritious food for enabling physical and mental health of school going students, to minimise the incidence of lifestyle diseases and their early onset in children. Further, it aimed to determine whether government schools in a district of New Delhi comply with the provisions of the Food Safety and Standards (Safe food and balanced diets for children in schools) Regulations, 2020 with respect to the major points in which the school may be directly responsible. The results show dismal compliance and implementation of a well intended regulation. The study has also provided recommendations which may be useful for informing policy in this urgent public health matter.

## Data Availability

All data produced in the present study are available upon reasonable request to the authors

## Declarations

The authors declare no conflict of interest. No funding has been received for this work. Data available at reasonable request to authors. Special thanks to Mr. Rajshekhar Pullabhatla and Group Captain P. Aanand Naidu of ISAC Centre for Built Environment Policy. The support from Mr Hardik Tomar for compilation of the data is gratefully acknowledged. The first author acknowledges Drexel Dornsife School of Public Health for the post doctoral fellowship.

## CREDiT Statement

Conceptualisation: RD, RS; Writing-Original Draft: RD, RS; Writing-review & editing: ALF, RD,RS; Supervision: ALF, RD; Validation: ALF, RD; Formal Analysis, Methodology and Project Administration: RS.

## Notes

### Competing Interest Statement

The authors have declared no competing interest.

### Funding Statement

This study did not receive any funding.

## References

1. ^a, b, c^Food Safety and Standards Authority of India. Food Safety and Standards (Safe food and balanced diets for children in school) Regulations 2020.

2. ^Government of India. Food Safety and Standards Act, 2006.

3. ^Food Safety and Standards Authority of India. (2019). FSSAI propose Ten-point charter for food sold, supplied to school children.

4. ^Raja Singh, Anil Dewan. (2022). Implementation of the Right to Healthy Environment: Regulations for running air conditioners in public buildings and recognition of biological pollutants. doi:10.21203/rs.3.rs-1270717/v1.

5. ^Viswanathan Mohan, Harish Ranjani, TS Mehreen, Rajendra Pradeepa, et al. (2016). Epidemiology of childhood overweight &amp; obesity in India: A systematic review. Indian J Med Res, vol. 143 (2), 160. doi:10.4103/0971-5916.180203.

6. ^Kaur S, Sachdev HP, Dwivedi SN, Lakshmy R, Kapil U.. (2008). Prevalence of overweight and obesity amongst school children in Delhi, India. Asia Pac J Clin Nutr., vol. 17.

7. ^Joel Fuhrman. (2018). The Hidden Dangers of Fast and Processed Food. American Journal of Lifestyle Medicine, vol. 12 (5), 375–381. doi:10.1177/1559827618766483.

8. ^Walter Willett, Johan Rockström, Brent Loken, Marco Springmann, et al. (2019). Food in the Anthropocene: the EAT–Lancet Commission on healthy diets from sustainable food systems. The Lancet, vol. 393 (10170), 447–492. doi:10.1016/s0140-6736(18)31788-4.

9. ^a, b^Hanieh-Sadat Ejtahed, Parham Mardi, Bahram Hejrani, Fatemeh Sadat Mahdavi, et al. (2024). Association between junk food consumption and mental health problems in adults: a systematic review and meta-analysis. BMC Psychiatry, vol. 24 (1). doi:10.1186/s12888-024-05889-8.

10. ^Maryam Tidjani Alou, Jean-Christophe Lagier, Didier Raoult. (2016). Diet influence on the gut microbiota and dysbiosis related to nutritional disorders. Human Microbiome Journal, vol. 1, 3–11. doi:10.1016/j.humic.2016.09.001.

11. ^Tina Rawal, Maartje Willeboordse, Monika Arora, Nitika Sharma, et al. (2021). Prevalence of Excessive Weight and Underweight and Its Associated Knowledge and Lifestyle Behaviors among Urban Private School-Going Adolescents in New Delhi. Nutrients, vol. 13 (9), 3296. doi:10.3390/nu13093296.

12. ^Maumita Kanjilal, Uma Kumar, Gajendra Kumar Gupta, Deepika Agrawal, et al. (2021). Dietary Habits and their Impact on the Physical Status of School Going Adolescents in Delhi: A Cross-sectional Study. JCDR. doi:10.7860/jcdr/2021/48202.15158.

13. ^Shalini Bassi, Deepika Bahl, Monika Arora, Fikru Tesfaye Tullu, et al. (2021). Food environment in and around schools and colleges of Delhi and National Capital Region (NCR) in India. BMC Public Health, vol. 21 (1). doi:10.1186/s12889-021-11778-6.

14. ^Neha Rathi, Lynn Riddell, Anthony Worsley. (2016). Food environment and policies in private schools in Kolkata, India: Table 1:. Health Promot. Int. doi:10.1093/heapro/daw053.

15. ^Vanitha B, Reddy R, Ranganath TS, Vishwanatha. (2019). Dietary Practices, Junk Food Consumption and Overweight among Adolescents in Schools of Urban Slums, Bengaluru. Natl J Community Med, vol. 10.

16. ^The Vantage Take. (2024). Why do schools sell junk food while asking children to avoid them?. First Post.

17. ^Samira Karnik, Amar Karnekar. (2012). Childhood Obesity: A Global Public Health Crisis. International Journal or Preventive Medicine, vol. 3.

18. ^Singh, Raja. (2020). RTI for Research: Using the Right to Information Act, 2005 for Research in India. Sandeep Kaur(BooksBonanza).

19. ^Pavitra Mohan, SanjanaBrahmawar Mohan, Manisha Dutta. (2019). Communicable or noncommunicable diseases? Building strong primary health care systems to address double burden of disease in India. J Family Med Prim Care, vol. 8 (2), 326. doi:10.4103/jfmpc.jfmpc_67_19.

20. ^Rukaiah Fatma Begum, Ankul Singh S, Sumithra Mohan. (2023). Impact of junk food on obesity and polycystic ovarian syndrome: Mechanisms and management strategies. Obesity Medicine, vol. 40, 100495. doi:10.1016/j.obmed.2023.100495.

21. ^Shriya Doreswamy, Anam Bashir, Jesus E Guarecuco, Simmy Lahori, et al. (2020). Effects of Diet, Nutrition, and Exercise in Children With Autism and Autism Spectrum Disorder: A Literature Review. doi:10.7759/cureus.12222.

22. ^Aleksandra Tomova, Igor Bukovsky, Emilie Rembert, Willy Yonas, et al. (2019). The Effects of Vegetarian and Vegan Diets on Gut Microbiota. Front. Nutr., vol. 6. doi:10.3389/fnut.2019.00047.

23. ^Ruo-Gu Xiong, Jiahui Li, Jin Cheng, Dan-Dan Zhou, et al. (2023). The Role of Gut Microbiota in Anxiety, Depression, and Other Mental Disorders as Well as the Protective Effects of Dietary Components. Nutrients, vol. 15 (14), 3258. doi:10.3390/nu15143258.

24. ^Tomas Hrncir. (2022). Gut Microbiota Dysbiosis: Triggers, Consequences, Diagnostic and Therapeutic Options. Microorganisms, vol. 10 (3), 578. doi:10.3390/microorganisms10030578.

25. ^Yi Zhang, Tatiana G. Kutateladze. (2018). Diet and the epigenome. Nat Commun, vol. 9 (1). doi:10.1038/s41467-018-05778-1.

26. ^Shammi Luhar, Dimple Kondal, Rebecca Jones, Ranjit M. Anjana, et al. (2020). Lifetime risk of diabetes in metropolitan cities in India. Diabetologia, vol. 64 (3), 521–529. doi:10.1007/s00125-020-05330-1.

27. ^Wei Perng, Rebecca Conway, Elizabeth Mayer-Davis, Dana Dabelea. (2023). Youth-Onset Type 2 Diabetes: The Epidemiology of an Awakening Epidemic. doi:10.2337/dci22-0046.

28. ^Dariush Mozaffarian. (2020). Dietary and policy priorities to reduce the global crises of obesity and diabetes. Nat Food, vol. 1 (1), 38–50. doi:10.1038/s43016-019-0013-1.

29. ^Lynnette M Neufeld, Eduardo B Andrade, Ahna Ballonoff Suleiman, Mary Barker, et al. (2022). Food choice in transition: adolescent autonomy, agency, and the food environment. The Lancet, vol. 399 (10320), 185–197. doi:10.1016/s0140-6736(21)01687-1.

30. ^Patricia Eustachio Colombo, Emma Patterson, Liselotte S Elinder, Anna Karin Lindroos. (2020). The importance of school lunches to the overall dietary intake of children in Sweden: a nationally representative study. Public Health Nutr., vol. 23 (10), 1705–1715. doi:10.1017/s1368980020000099.

31. ^World Health Organization. (2022). “Food marketing exposure and power and their associations with food-related attitudes, beliefs and behaviours: a narrative review.”.

32. ^Viswanathan Mohan, Harish Ranjani, Rajendra Pradeepa, TS Mehreen, et al. (2014). Determinants, consequences and prevention of childhood overweight and obesity: An Indian context. Indian J Endocr Metab, vol. 18 (7), 17. doi:10.4103/2230-8210.145049.

33. ^Viswanathan Mohan, Harish Ranjani, Rajendra Pradeepa, TS Mehreen, et al. (2014). Determinants, consequences and prevention of childhood overweight and obesity: An Indian context. Indian J Endocr Metab, vol. 18 (7), 17. doi:10.4103/2230-8210.145049.

34. (2024). Few takers for ‘healthy campuses’ initiated by FSSAI and UGC. Education Times.

35. ^Directorate of Education: School Branch. (2020). Circular titled ‘Regarding safe food and healthy diets for school children’. Government of NCT of Delhi.

36. ^Surabhi Gautam, Romsha Kumar, Uma Kumar, Sanjeev Kumar, et al. (2023). Yoga maintains Th17/Treg cell homeostasis and reduces the rate of T cell aging in rheumatoid arthritis: a randomized controlled trial. Sci Rep, vol. 13 (1). doi:10.1038/s41598-023-42231-w.

37. ^Mrithunjay Rathore, Meghnath Verma, Jessy Abraham, Rima Dada, et al. (2023). Impact of “Yoga Based Lifestyle Interventions” and its Implications on Health and Disease. IRJAY, vol. 06 (01), 64–73. doi:10.47223/irjay.2023.6112.

38. ^Madhuri Tolahunase, Rajesh Sagar, Rima Dada. (2017). Impact of Yoga and Meditation on Cellular Aging in Apparently Healthy Individuals: A Prospective, OpenLLabel SingleLArm Exploratory Study. Oxidative Medicine and Cellular Longevity, vol. 2017 (1). doi:10.1155/2017/7928981.

39. ^Rima Dada, Surabhi Gautam, Tanuj Dada, Prabhakar Tiwari, et al. (2023). Yoga: unraveling the internal pharmacy – Impact on genome and epigenome. MRAJ, vol. 11 (12). doi:10.18103/mra.v11i12.4877.

40. ^K. Sujatha Rao. (2017). Do We Care?. doi:10.1093/acprof:oso/9780199469543.001.0001.

41. ^Raja Singh. (2023). Signboards prohibiting tobacco sale within 100 yards of educational institutes: the appraisal of prohibition compliance and on-ground status of the anti-smoking law in New Delhi’s major administrative precinct. Cities &amp; Health, vol. 7 (5), 719–728. doi:10.1080/23748834.2023.2215417.

42. ^Sani Dimitroulopoulou, Marzenna R. Dudzińska, Lars Gunnarsen, Linda Hägerhed, et al. (2023). Indoor air quality guidelines from across the world: An appraisal considering energy saving, health, productivity, and comfort. Environment International, vol. 178, 108127. doi:10.1016/j.envint.2023.108127.

43. ^Raja Singh, Arthur L Frank. (2023). Does the Presence of Asbestos-Containing Materials in Buildings Post-construction and Before Demolition Have an Impact on the Exposure to Occupants in Non-occupational Settings?. doi:10.7759/cureus.37305.

